## Supplemental File 1 COREQ checklist for "Implementation and enforcement of mandatory calorie labelling regulations for the out-of-home sector in England: qualitative study of the experiences of business implementers and regulatory enforcers"

**Supplementary File 1: COREQ checklist (32 items)**

| **Domain 1: Research team and reflexivity** | **Answer** | **Location in manuscript (Section, page no.)** |
| --- | --- | --- |
| **Personal Characteristics** |  |  |
| 1. Interviewer/facilitator  Which author/s conducted the interview or focus group? | ME | Methods,  Pages 4,5 |
| 2. Credentials  What were the researcher’s credentials? | All authors title page. Interviewer’s elaborated page 5. | Title page, specifics on page 5 Reflexivity |
| 3. Occupation  What was their occupation at the time of the study? | Yes | Methods,  Page 5 Reflexivity |
| 4. Gender  Was the researcher male or female | Yes | Methods,  Page 5 Reflexivity |
| 5. Experience and training  What experience or training did the researcher have? | Yes | Methods,  Page 5 Reflexivity |
| **Relationship with participants** |  |  |
| 6. Relationship established  Was a relationship established prior to study commencement? | Only for the purposes of this research | n/a |
| 7. Participant knowledge of the interviewer  What did the participants know about the researcher? e.g. personal goals, reasons for doing the research | All contacts were sent a Participant Information Sheet (Supplementary File 2) explaining the study and its purpose. After the introductory call, if the potential participants met the inclusion criteria and were willing to proceed, then they signed an electronic consent form prior to the recorded interview. | Methods Page 4 & Participant Information Sheet (Suppl. File 2) |
| 8. Interviewer characteristics  What characteristics were reported about the interviewer/facilitator? e.g. Bias, assumptions, reasons and interests in the research topic | Yes | Methods,  Page 5 Reflexivity |
| **Domain 2: study design** |  |  |
| **Theoretical framework** |  |  |
| 9. Methodological orientation and Theory  What methodological orientation was stated to underpin the study?  e.g. grounded theory, discourse analysis, ethnography, phenomenology, content analysis | Framework Method | Methods,  Page 4 |
| **Participant selection** |  |  |
| 10. Sampling  How were participants selected?  e.g. purposive, convenience, consecutive, snowball | Purposive | Methods,  Page 3 |
| 11. Method of approach  How were participants approached?  e.g. face-to-face, telephone, mail, email | Yes | Methods,  Pages 3,4 |
| 12. Sample size  How many participants were in the study | 20 | Results,  Page 5 |
| 13. Non-participation  How many people refused to participate or dropped out? Reasons? | Included | Methods,  Pages 3-4 |
| **Setting** |  |  |
| 14. Setting of data collection  Where was the data collected? e.g. home, clinic, workplace | Video conference | Methods,  Page 4 |
| 15. Presence of non-participants  Was anyone else present besides the participants and researchers? | No | - |
| 16. Description of sample  What are the important characteristics of the sample? e.g. demographic data, date | Included | Table 1, Results,  Page 6 |
| **Data collection** |  |  |
| 17. Interview guide  Were questions, prompts, guides provided by the authors? Was it pilot tested? | Given the aim to evaluate and inform policy, the Framework method was used for data analysis with specific questions regarding barriers and facilitators to implementation and enforcement. | Methods,  Page 4 |
| 18. Repeat interviews  Were repeat interviews carried out?  If yes, how many | No | - |
| 19. Audio/visual recording  Did the research use audio or visual recording to collect the data? | Interviews were audio-recorded with participants’ consent. | Methods,  Page 4 |
| 20. Field notes  Were field notes made during and/or after the interview or focus group? | Yes | Methods,  Page 3 |
| 21. Duration  What was the duration of the interviews or focus group? | Interviews lasted for a maximum of 60 minutes. | Methods,  Page 4 |
| 22. Data saturation  Was data saturation discussed? | Yes | Methods,  Pages 3,4 |
| 23. Transcripts returned  Were transcripts returned to participants for comment and/or correction? | Interviews were digitally audio recorded and transcribed verbatim by a professional transcription company. Transcripts were checked for accuracy. Transcripts were not returned to participants for comment or correction. | Methods,  Page 4 |
| **Domain 3: analysis and findings** |  |  |
| **Data analysis** |  |  |
| 24. Number of data coders  How many data coders coded the data? | Three researchers (ME, JA, MW) independently coded three transcripts to ensure important aspects of the data were not missed, and ME coded all remaining transcripts. | Methods,  Page 4 |
| 25. Description of the coding tree  Did authors provide a description of the coding tree? | Five key themes, including their more detailed descriptions are presented in Table 2 | Results,  Page 7,  Table 2 |
| 26. Derivation of themes  Were themes identified in advance or derived from the data | Included | Methods,  Page 4 |
| 27. Software  What software, if applicable, was used to manage the data? | NVivo, version 12 | Methods,  Page 5 |
| 28. Participant checking  Did participants provide feedback on the findings? | No | n/a |
| **Reporting** |  |  |
| 29. Quotations presented  Were participant quotations presented to illustrate the themes/findings?  Was each quotation identified?  e.g. participant number | Yes, quotations were presented and attributed to anonymised participants. | Results,  Pages 8-14 |
| 30. Data and findings consistent  Was there consistency between the data presented and the findings? | Yes | Results,  Pages 8-14 |
| 31. Clarity of major themes  Were major themes clearly presented in the findings? | Yes | Results,  Pages 8-14 |
| 32. Clarity of minor themes  Is there a description of diverse cases or discussion of minor themes? | Yes | Results,  Pages 8-14 |
