## Supplemental File 2 Participant Information Sheet for "Implementation and enforcement of mandatory calorie labelling regulations for the out-of-home sector in England: qualitative study of the experiences of business implementers and regulatory enforcers"

#### Summary

In April 2022, the UK Government's Department of Health & Social Care introduced a requirement for large chain businesses in the out of home food sector to present calorie labels on their menus.

We want to understand the experiences of those involved with making menu labelling happen in large out-of-home food businesses and those involved in enforcing the regulations.

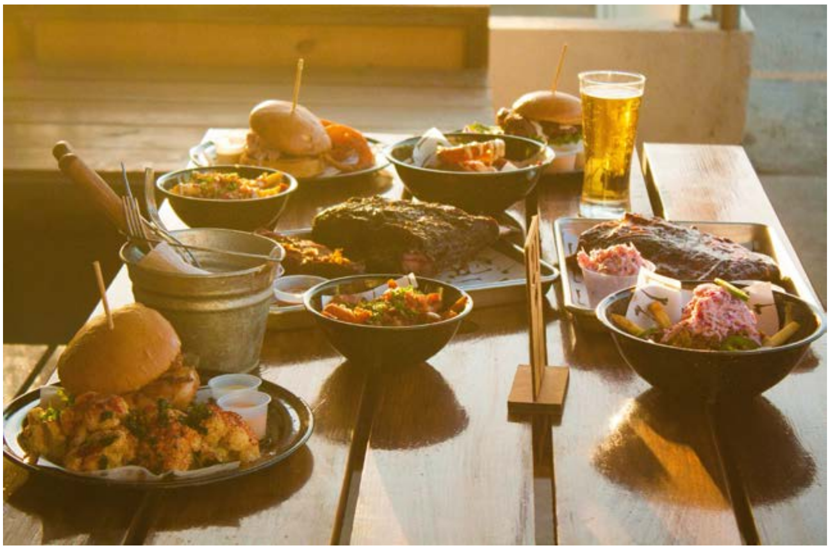

Photo by Marco Guerrero on Unsplash.

Please take the time to read the following information carefully. Discuss it with colleagues if you wish and decide whether you wish to take part. Thank you for your support with our research so far.

#### Contents

1 Why we are doing this study
2 Why am I being asked to take part?
3 What will happen to me if I take part?
4 Possible benefits and disadvantages of taking part
5 More information about taking part
6 Contact for further information

#### How to contact us

If you have any questions about this study, then please talk to:

**Dr Michael Essman**
MRC Epidemiology Unit
University of Cambridge School of Clinical Medicine
Box 285 Institute of Metabolic Science
Cambridge Biomedical Campus
Cambridge CB2 0QQ


#### 1 Why we are doing this study?

##### What are we studying?

In this research we are aiming to interview people responsible for implementing and enforcing the menu labelling regulations. We will ask whether the rules did what they were supposed to, and if there were any problems with them. We will share our results with people working in Government, other researchers, and the public. Our results could help Government refine calorie labelling rules.

#### 2 Why am I being asked to take part?

We are inviting you to take part because you work for an organisation responsible for either implementing the menu labelling regulations at a large out-of-home food business or an organisation responsible for enforcing the menu labelling regulations.

#### 3 What will happen to me if I take part?

We will conduct **one-to-one interviews via Zoom** with individuals working with large out-of-home food businesses that are working to implement the menu labelling regulations. Participants will be asked to only complete one interview, and each interview will last up to 60 minutes.

During each interview, we will aim to better understanding your experience with implementing or enforcing the menu labelling regulations, including the implementation process; the financial impacts of intervention implementation; enforcement activity and its success; barriers and enablers to enforcement; and the financial costs of enforcement.

Before taking part in an interview, you will have the opportunity to speak with a researcher and ask questions about the study. If you are willing to take part, you will be asked to provide consent to take part using an online form (e-consent).

The interviews will be audio-recorded to help us retain key insights shared. No one will have access to the recording except members of the research team at the University of Cambridge.

#### 4 Possible benefits and disadvantages of taking part

##### What are the possible benefits of taking part?

You will help us understand the impacts of the menu labelling regulations from people working on your side of implementation or enforcement. You will have the opportunity to share your perspectives that can provide feedback and research data for the UK Government's Department of Health and Social Care.

##### What are the possible disadvantages and risks of taking part?

Taking part in this study will involve sacrificing some of your time.

### 5 More information about taking part

#### Do I have to take part?

No, it is up to you to decide whether to take part. You are free to withdraw at any time, without giving a reason.

#### Will I receive any payment for taking part?

You will not receive any form of payment for taking part in this study.

#### What if there is a problem?

If you have a concern about any aspect of this study, you should ask to speak to the research team who will do their best to answer your questions. If you remain unhappy and wish to complain formally, the University of Cambridge complaints process is available to you through the University of Cambridge Clinical School Secretary: telephone: 01223 333543 or

#### What will happen to information about me collected during the study?

The interviews will be audio-recorded; the recording will be sent to an external, professional transcription team to be typed up. The transcription team will remove all names from the transcript, and it will only be identified by an anonymised code. Information transferred between us and the transcription team will be subject to the same high standard of data security and confidentially for all our research data. Apart from the transcription team, no one will have access to the recording except members of the research team at the University of Cambridge.

Information we collect during the research will be kept strictly confidential. Any information about you will have your name and affiliation removed so that you cannot be recognised from it and it will not be used or made available for any purpose other than for research. There may be anonymised quotations from the interviews within any reports or publications.

With your permission, information will be stored anonymously at the MRC Epidemiology Unit on a secure research drive. Codes connecting your individual identity to the stored data records will be kept separately. The database containing personal information is on a secured network drive on computers in the MRC Epidemiology Unit, University of Cambridge.

Occasionally our studies may be monitored by our Sponsors. This is to ensure our research is conducted soundly. This procedure is routine and carried out by fully qualified personnel and data confidentiality will be always adhered to. At the end of the study the confidential records will be kept for a minimum of 20 years and then destroyed.

The University of Cambridge is the sponsor for this study based in the United Kingdom. We will be using information from you to undertake this study and will act as the data controller for this. Cambridge University will keep identifiable information about you for a minimum of 20 years after the study has finished.

Your rights to access, change or move your information are limited, as we need to manage your information in specific ways for the research to be reliable and accurate. If you withdraw from the study, we will keep the information about you that we have already obtained. To safeguard your rights, we will use the minimum personally identifiable information possible.

You can find out more about how we use your information at https://www.medschl.cam.ac.uk/research/privacy-notice-how-we-use-your-research-data/

#### What will happen to the results of the study?

When the study is completed, the results will be published in an academic journal without a paywall so that anyone can see the results. We may also present the results orally at scientific meetings and to interested stakeholders, including representatives from the UK Government's Department of Health and Social Care. If published or presented, your identity, affiliation and personal details will be kept confidential. No information that could identify you, like your name, will be published in any report about this study. We will also send you a copy of any publications.

#### Who is organising and funding the study?

This study is organised by the Centre for Diet and Activity Research, which is in the MRC Epidemiology Unit, part of the University of Cambridge. The study is funded by the National Institute for Health Research and is also receiving support from the MRC Epidemiology Unit.

#### Who has reviewed the study?

This trial has been reviewed by an independent group of people, called a Research Ethics Committee, to protect your safety, rights, wellbeing, and dignity. The study has been granted approval by the University of Cambridge Humanities and Social Sciences Research Ethics Committee. You can contact the Committee by quoting the ethics reference 22.294 by

### 6 Contact for further information

If you have any questions regarding the study or how you might be involved further contact information can be found below.

#### Study Lead

**Dr Michael Essman**
Research Associate
MRC Epidemiology Unit
University of Cambridge


#### Principal Investigator

**Dr Jean Adams**
Programme Leader and MRC Investigator
MRC Epidemiology Unit
University of Cambridge


#### Thank you for taking the time to consider taking part in this study.
