## Supplemental File 3 Interview Topic Guide Implementers for "Implementation and enforcement of mandatory calorie labelling regulations for the out-of-home sector in England: qualitative study of the experiences of business implementers and regulatory enforcers"

**Data collection for WP5: Interviews with intervention implementers**

**Interview topic guide**

*Note: use separate topic guide for each interview.*

**Interview #:______________**

**Pre-interview checklist for interviewer:**

**☐** Participant information sheet reviewed by interviewee

**☐** Consent process completed and uploaded to SRD

Interview conducted via:

**☐** telephone	OR **☐** Zoom

**Participant information**

**Introduction**

Thanks for agreeing to take part in an interview for our study today. This interview is expected to take up to a maximum of 60 minutes. Is that OK with you, and do you need to finish earlier for any reason?

My name is Mike Essman, I work with the Centre for Diet and Activity Research at Cambridge University.

Before we start on the interview, I’m going to briefly remind you of the policy we are going to discuss today. In April 2022, England’s Department of Health & Social Care introduced a requirement for large chain businesses in the out of home food sector to present calorie labels on their menus.

We want to understand the experiences of those involved with making menu labelling happen in large out of home food businesses. As a reminder, all questions are optional – we can skip any questions that you do not feel comfortable answering.

Are you happy that you understand the research project and what it will entail? [*Let them answer]*

Do you have any questions before we start? [*Let them answer]*

I will now switch on the audio recorder.

[SWITCH ON AUDIO RECORDER]

*Discussion begins (below), make sure to give people time to think and don’t move too quickly. Use the probes to make sure that all issues are addressed, but move on when you feel you are starting to hear repetitive information and allow for flexibility in the direction that the participant takes the conversation.*

| **Check** | **#** | **Question** | **Probe** |
| --- | --- | --- | --- |
| **Introduction – ice breakers and overall understanding of regulations** | | | |
| ☐ | 1 | Firstly, could tell me what your professional role is? | -What organisation do you work for? Follow up asking the type of organisation, how do they describe themselves. What type of food do you serve?  -How long have you worked there?  -What role do you fulfil within this organisation?  -What are your main priorities? |
| ☐ | 2 | What do you understand about the menu labelling regulations? (can use any other terminology the interviewee uses for the policy) | -What business types are subject to the regulations?  -What food is covered?  -What info is supposed to be presented and where? (answer: kcal at point of sale)  -Are there any exemptions?  (*Use my notes to prompt and understand how much they cover. After they answer, I can use my notes to make sure they clearly understand the law. If anything seems unclear, share PowerPoint slide to check understanding)* |
| ☐ | 3 | *[If not answered in #1,2*] Before this interview, what (if any) involvement have you had in the labelling regulations? | How have you been involved in the menu labelling policy? And its regulation? |
| **Policy Implementation – feasibility, barriers, and efficacy** | | | |
| We are interested in your experience with implementing the menu labelling policy, and the costs and benefits associated with implementing the policy. I want to emphasise that I am not concerned about assessing your level of compliance, but instead want to know how you feel your organisation has been impacted by the menu labelling policy. | | | |
| ☐ | 4 | What has implementing the menu labelling requirements involved for you and/or your organisation? | -For background, when and how did your organisation start making calorie content information available ie were they already doing it before the regulations were introduced?  - How did you implement it? For example, how to you make sure (1) that labels are present, (2) that labels are displayed correctly, and (3) that labels are accurate?  -How did you calculate calorie content, and why that way?  -Are there other ways you could’ve chosen to implement and calculate kcal? Why not them?  -How accurate do you think your calorie labelling is (i.e. labels reflect actual energy content)? Do you have a way of testing the label accuracy? Do you think other chains will be providing accurate calorie labels, and are you working with any companies on the regulations?  -How did you decide how to design and display calorie labels, and why that way?  -Are there other ways you could’ve chosen to label kcal? Why not them?  -What has been your policy on online ordering or other forms of food orders? |
| ☐ | 5 | How has the process of implementing the menu labelling requirements gone for you/your organisation? | -What has gone well?  -What has gone not so well? -What has helped?  -What has hindered?  -Is there been anything that you’ve particularly struggled with?  -Is there anything you’d say was just impossible or infeasible to do? |
| ☐ | 6 | What impacts has implementing the menu labelling requirements had on your organisation? | -Have you changed your products, including reformulation, because of your calorie assessments/labelling process or customer responses to the regulations? (reduced portion size, more healthy choices, reformulation, change in cooking methods, new products introduced)  - Has the company approach to the development and introduction of new products changed since the introduction of the regulations? If so, how?  -Has there been an admin burden, and if so, what?  -Have there been any effects on your staff including how you train them or otherwise? Do staff see the regulations as positive or negative?  -Has it cost the organisation money or other resources, and do you know how much?  -Has it led to strategy/processes changing in other ways? For example, is there is anything your company hasn’t been able to do because you’ve been implementing this, or anything else you’ve had to do as a follow up? (interested in ‘compensatory practices’)  -Have you received any feedback from customers about the menu labelling?  -Have there been any other impacts that could be helpful for us to understand? |
| ☐ | 7 | How are you and your organisation expecting enforcement to occur? | -For those tasked with enforcing the menu labelling requirements, how do you think they will verify accuracy of menu labels? Do you think it will be possible to enforce the requirements?  -Do you think their enforcement also comes with costs? |
| **Summing up – Overall effects of the policy** | | | |
| Thank you for your responses so far. Finally, we are interested in some of your overall thoughts about the effects of the menu labelling requirements. | | | |
| ☐ | 8 | Do you think the menu labelling requirements will “work”? | -How are the regulations perceived within the organisation?  Are they seen as a threat or an opportunity?  -Does the company monitor the impact?  What has it learned about impact on consumer choice?  What direct feedback has it received from customers?  -Has the guidance issued by DHSC been helpful, was there anything missing, what additional support would you like to see? |
| ☐ | 9 | Do you think the menu labelling requirements will “work”? | -Explore what they think “work” means in terms of government policy goals; reasons why/why not.  -Do you think everyone will adhere to the rules?  -What impact do you think it might have on consumers? |
| ☐ | 10 | What do you view as the main problem(s) that the proposed menu labelling requirements are trying to address? (*if not captured*) | Probe for evidence: help consumers choose, health improvements, transparency of food offerings |
| **Concluding remarks** | | | |
| ☐ | 11 | That’s all the questions I have for you today, thank you again for your contribution. *Briefly review discussion, ask any questions I missed or didn’t get full answers to.*  Do you have any questions, or is there anything we haven’t covered which you think might be important in our research before I switch off the recorder? |  |
| ☐ | 12 | Also, do you have colleagues in your organisation or elsewhere that you would recommend we speak to? *Read inclusion criteria to them.* | ****Add names and contact details to longlist after interview**** |
| Thank you for your time. I’m going to switch off the recorder now. [SWITCH OFF AUDIO RECORDER] | | | |
| **Data collection diary** | | | |
| *Add anonymous comments only, to aid interpretation of transcripts – store in SRD* | | | |

**Post-interview checklist for interviewer:**

**☐** Audio recording uploaded to SRD and checked that the voices are clear, etc.

**☐** Transcript request form completed and sent to Inge Loudon.

**☐** Audio recording deleted from recording device

**☐** *If applicable* add contact(s) offered for snowball sampling to the interview contact list

****INTERVIEWER MUST UPLOAD THIS DOCUMENT TO SRD UPON COMPLETION****
