## Supplemental File 4 Interview Topic Guide Enforcers for "Implementation and enforcement of mandatory calorie labelling regulations for the out-of-home sector in England: qualitative study of the experiences of business implementers and regulatory enforcers"

**Data collection for WP5: Interviews with intervention enforcers**

**Interview topic guide**

*Note: use separate topic guide for each interview.*

**Interview #:______________**

**Pre-interview checklist for interviewer:**

**☐** Participant information sheet reviewed by interviewee

**☐** Consent process completed and uploaded to SRD

Interview conducted via:

**☐** telephone	OR **☐** Zoom

**Participant information**

**Introduction**

Thanks for agreeing to take part in an interview for our study today. This interview is expected to take up to a maximum of 60 minutes. Is that OK with you, and do you need to finish earlier for any reason?

My name is Mike Essman, I work with the Centre for Diet and Activity Research at Cambridge University.

Before we start on the interview, I’m going to briefly remind you of the policy we are going to discuss today. In April 2022, England’s Department of Health & Social Care introduced a requirement for large chain businesses in the out of home food sector to present calorie labels on their menus.

We want to understand the experiences of those involved with enforcing the menu labelling regulations in effect for large out of home food businesses. As a reminder, all questions are optional – we can skip any questions that you do not feel comfortable answering.

Are you happy that you understand the research project and what it will entail? [*Let them answer]*

Do you have any questions before we start? [*Let them answer]*

I will now switch on the audio recorder.

[SWITCH ON AUDIO RECORDER]

*Discussion begins (below), make sure to give people time to think and don’t move too quickly. Use the probes to make sure that all issues are addressed, but move on when you feel you are starting to hear repetitive information and allow for flexibility in the direction that the participant takes the conversation.*

| **Check** | **#** | **Question** | **Probe** |
| --- | --- | --- | --- |
| **Introduction – ice breakers and overall understanding of regulations** | | | |
| ☐ | 1 | Firstly, could tell me what your professional role is? | -What organisation do you work for?  -How long have you worked there?  -What role do you fulfil within this organisation?  -What are your main priorities? |
| ☐ | 2 | What do you understand about the menu labelling regulations? (can use any alterative terminology the interviewee uses for the policy) | -What business types are subject to the regulations? Do you know how many there are in your district/area? How many staff are responsible for enforcement?  -What food is covered?  -What info is supposed to be presented and where? (answer: kcal at point of sale)  -Are there any exemptions?  (*Use my notes to prompt and understand how much they cover. After they answer, I can use my notes to make sure they clearly understand the law. If anything seems unclear, share PowerPoint slide to check understanding)* |
| **Policy Implementation – feasibility, barriers, and efficacy** | | | |
| We are interested in your experience with implementing the menu labelling policy, and the costs and benefits associated with implementing the policy. | | | |
| ☐ | 4 | What has enforcing the menu labelling requirements involved for you and/or your organization? | -What do you or your organization mean by enforcement?  - How do you identify businesses for inspection?  - Have you received and responded to complaints from members of the public?  - How easy/difficult is it to identify businesses in scope?  - What does your enforcement/inspection process look like? For example, are you checking (1) if labels are present, (2) if labels are displayed correctly, and (3) if they are accurate?  -How accurate do you think the calorie labelling is (i.e. labels reflect actual energy content)? Do you have a way of checking the label accuracy?  -Are you able to verify the accuracy of menu labels, and if so, how? Do you feel confident in your ability to enforce the requirements?    -Are there other ways you could’ve chosen to verify the accuracy of calorie labels, and why didn’t you choose them? Some examples could be sending meals to labs, looking if labels seem plausible, asking for paperwork, etc.  - How well-informed are local businesses about the regulations? Have you played a role in this?  -Do you think most chains are providing accurate calorie labels, and do you know if any chains are working together on implementation?  -What has been your policy on online ordering or other forms of food orders? |
| ☐ | 5 | How has the process of enforcing the menu labelling requirements gone for you/your organisation? | -What has gone well?  -What has gone not so well? -What has helped?  -What has hindered?  -Is there been anything that you’ve particularly struggled with?  -Is there anything you’d say was just impossible or infeasible to do? |
| ☐ | 6 | What impacts have enforcing the menu labelling requirements had on your organisation? | -What are the costs of enforcement to your organisation? (administrative burden, enforcement staff time, other costs?)  -Has it led to strategy/processes changing in other ways? For example, is there is anything your company hasn’t been able to do because you’ve been implementing this, or anything else you’ve had to do as a follow up? What is the priority of OOH calorie labelling compared with other work areas? How do you balance the workload across different work within your organisation? (interested in ‘compensatory practices’)  -Have there been any other impacts that could be helpful for us to understand?  What further information would you find helpful to successfully enforce the regulations? |
| ☐ | 7 | I’m interested in some of the specific of how outlets have complied with the menu labelling requirements. Overall, how do you think compliance is going at the outlets you’ve examined? | Overall, would you say that…  -Are most outlets providing labelling? Are labels provided for all food items? Non-alcoholic drink items?  -Is calorie labelling provided per portion?  -Is calorie information and reference information displayed clearly and prominently? |
| **Summing up – Overall effects of the policy** | | | |
| Thank you for your responses so far. Finally, we are interested in some of your overall thoughts about the effects of the menu labelling requirements. | | | |
| ☐ | 8 | Do you think the menu labelling regulations will achieve their goals? | Explore what they think “work” means in terms of policy goals; reasons why/why not.  -Do you think everyone will adhere to the rules?  -What impact do you think it might have on consumers? |
| ☐ | 9 | Do you think the proposed menu labelling requirements will affect anything else? | e.g., reformulation of options, new/removed options, increase/decrease in prices, changes in meal/portion sizes |
| ☐ | 10 | What do you view as the main problem(s) that the proposed menu labelling requirements are trying to address? (*if not captured*) | Probe for evidence: help consumers choose, health improvements, transparency of food offerings |
| **Concluding remarks** | | | |
| ☐ | 11 | That’s all the questions I have for you today, thank you again for your contribution. *Briefly review discussion, ask any questions I missed or didn’t get full answers to.*  Do you have any questions, or is there anything we haven’t covered which you think might be important in our research before I switch off the recorder? |  |
| ☐ | 12 | Also, do you have colleagues in your organisation or elsewhere that you would recommend we speak to? *Read inclusion criteria to them.* | ****Add names and contact details to longlist after interview**** |
| Thank you for your time. I’m going to switch off the recorder now. [SWITCH OFF AUDIO RECORDER] | | | |
| **Data collection diary** | | | |
| *Add anonymous comments only, to aid interpretation of transcripts – store in SRD* | | | |

**Post-interview checklist for interviewer:**

**☐** Audio recording uploaded to SRD and checked that the voices are clear, etc.

**☐** Transcript request form completed and sent to Inge Loudon.

**☐** Audio recording deleted from recording device

**☐** *If applicable* add contact(s) offered for snowball sampling to the interview contact list

****INTERVIEWER MUST UPLOAD THIS DOCUMENT TO SRD UPON COMPLETION****
